# Modeling the HIV epidemic in MSM in Cyprus: Reaching only the 95-95-95 cascade of care targets fails to reduce HIV incidence by 90% in 2030

**DOI:** 10.1101/2023.01.29.23285158

**Authors:** I. Gountas, K. Pantavou, G. Siakalis, A. Demetriou, I. Demetriades, G. K. Nikolopoulos

## Abstract

**Objectives:** National responses should be improved and accelerated to meet the target of ending the Acquired ImmunoDeficiency Syndrome (AIDS) epidemic by 2030. In the Republic of Cyprus, Men who have Sex with Men (MSM) are disproportionately affected by Human Immunodeficiency Virus (HIV), accounting approximately for half of all annual HIV diagnoses. This study aims to assess the evolution of HIV incidence in MSM in Cyprus until 2030 under different scenarios.

**Methods:** A model of HIV transmission and progression was calibrated to Cypriot epidemiological data. Four scenarios were examined: status quo, two scenarios focusing on introducing Pre-Exposure Prophylaxis (PrEP), and a 90% HIV incidence reduction scenario.

**Results:** Reaching only the 95-95-95 HIV cascade of care (CoC) targets among MSM fails to achieve the 90% reduction in HIV incidence: the estimated reduction in 2030 compared to 2015 would be 48.6%. Initiating a PrEP intervention only for high risk MSM would cause a modest further reduction in HIV incidence. To meet the 90% HIV reduction target, PrEP should be expanded to both high and medium risk MSM and, after 2025, behavioral interventions should be implemented so as high risk MSM gradually move to the medium risk category.

**Conclusions:** Cyprus will not reach the HIV incidence reduction target by 2030 unless PrEP is gradually promoted and delivered to all high and medium risk MSM along with awareness and behavioral interventions.

## Introduction

The third Sustainable Development Goal (SDG-3) includes, as a specific target, the end of Acquired ImmunoDeficiency Syndrome (AIDS) epidemic by 2030 ^1,2^. In 2016, the Political Declaration of the United Nations (UN) General Assembly on Human Immunodeficiency Virus (HIV) and AIDS committed countries to a fast-track strategy released by the Joint United Nations Programme on HIV and AIDS (UNAIDS) in 2014. That strategy recommended that, by 2020, 90% of people living with HIV (PLHIV) should know their HIV status, 90% of people who know their status should be on antiretroviral treatment (ART), and 90% of people on ART should achieve viral suppression [i.e., 90-90-90 cascade of care (CoC) targets] ^2,3^. According to mathematical modelling, if the 90-90-90 CoC targets were reached in 2020, with ART coverage and viral suppression rising to 95% in 2030, HIV incidence was estimated to decrease by 90% in 2030, the year of ending the AIDS epidemic ^3,4^.

Despite considerable progress, the world fell short of the 2020 CoC targets ^5–7^. Modeling studies at the country level underline that, in high-income countries, even the 95-95-95 CoC targets do not suffice to reduce HIV incidence by 2030 ^8,9^. To meet the HIV incidence reduction target, additional interventions including condom distribution, programs for behavior change, and delivery of Pre-Exposure Prophylaxis (PrEP) should be implemented ^10^.

In the Republic of Cyprus, MSM is the most prevalent HIV group, representing approximately half of all annual HIV diagnoses ^11^. The 2020 CoC targets in this population were practically reached; 92% of MSM living with HIV were diagnosed, 88% of those diagnosed were on ART, and 93% of those on ART were virally suppressed (Table S1). However, it is unknown whether Cyprus is on track to meet the 2030 HIV incidence reduction target of 90%. Additionally, although PrEP is an effective and safe prevention measure ^12,13^, it is currently unavailable in Cyprus and its expected effect has not yet been quantified.

The aim of this study was: i) to estimate the expected reduction in HIV incidence by 2030 under a status quo scenario (without PrEP); ii) to quantify the effect of introducing PrEP, and iii) to describe the required interventions in order to meet the target of 90% reduction in HIV incidence by 2030. The findings of this study could facilitate the national HIV/AIDS response and the forthcoming discussions about PrEP introduction and delivery in Cyprus.

## Methods

### Description of the HIV transmission model

A discrete-time, stochastic, dynamic, individual-based model of HIV transmission and progression, and of the CoC ^14^ was fitted to epidemiological data from the Cypriot epidemic (Table 1). The structure of the model is shown in detail in Figure S1 in the supplementary material. The model describes the transitions between the following stages: (a) susceptible MSM, (b) HIV-infected MSM who are not linked to care, (c) HIV-infected MSM who are ART without viral suppression, and (d) HIV-infected MSM on ART with viral suppression. After infection, MSM progress to the undiagnosed stage. Due to higher viremia levels, people in the recent phase of their HIV infection have a higher probability of HIV transmission than those who are longer-term infected ^15,16^. Every year, a time-dependent number of MSM are diagnosed and progress to the “infected and diagnosed” compartment of the model. When MSM are diagnosed, they can later progress to the ART compartment. ART can lead to virological suppression with a certain probability. HIV-infected people with viral suppression are less likely to transmit HIV than those who have not achieved virological response. It is assumed that the virological response reduces the risk of sexual transmission of HIV by 93% ^15,16^ (Τable 1). HIV-infected MSM can move back to the “infected and diagnosed” compartment of the model due to loss of follow-up. To model heterogeneity, the population was additionally stratified by risk behavior group according to data from the European MSM Internet Survey 2017 (EMIS-2017) ^17^: high risk, medium risk, and low risk. The model assumes that the proportion of high/medium/low remains constant in MSM until the last year of the projections. We assumed that only MSM belonging to the high or medium risk group could be infected. The model allowed MSM to move between risk states.

**Table 1.** Model parameters, input values, and sources.

| Parameter | Value | Source |
| --- | --- | --- |
| Estimated number of MSM | 6,954 | Marcus et al. <sup>21</sup> |
| Estimated number of people living with HIV and CoC | Table S1<sup>a</sup> | MoH of Cyprus and Grigorios Clinic (the reference HIV clinic in Cyprus) |
| MSM risk behavior | Table S2<sup>a</sup> | EMIS-2017 Cyprus data <sup>19</sup> |
| Movement between risk groups | 2 years | O Murchu et al. <sup>22</sup> |
| Reduction in transmission rate while on ART | 93% | Donnell et al. <sup>16</sup> |
| Effectiveness of PrEP | 86% | McCormack et al. <sup>23</sup> |
| Co-factor increase in HIV transmission probability during the initial acute phase with high viremia | 14.5 | Borquez et al. <sup>15</sup> |

<sup>a</sup> Table in the supplementary material.
Abbreviations: ART, antiretroviral treatment; CoC, cascade of care; EMIS, European MSM Internet Survey; HIV, Human Immunodeficiency Virus; MoH, Ministry of Health; MSM, Men who have Sex with Men; PrEP, Pre-Exposure Prophylaxis.

### Model calibration and validation

The model was calibrated to match the trajectory of the Cyprus HIV CoC between 2014 and 2020. The total number of HIV infections in 2014-2020 was estimated using the HIV ECDC (European Centre for Disease Prevention and Control) tool ^18^. The numbers of HIV diagnoses and of MSM who were on ART were retrieved from the Grigorios clinic (the reference HIV clinic in the Republic of Cyprus). The infection rate and the probabilities of HIV diagnosis, ART initiation, and viral suppression varied until the model reproduced the observed HIV epidemic in Cyprus.

Each scenario included 1000 runs. The median and 2.5th/97.5th percentiles of the simulations are shown to address uncertainty in model projections (stochastic variability). More details about the calibration procedure and the data used for calibration are available in the supplementary material.

### Modelling scenarios

Four scenarios were examined:

1. Status quo scenario: The status quo scenario was used to generate predictions regarding the current HIV strategy (without PrEP).
2. Scenario 2 (S2): Launching an intervention in 2023 so as all of the high risk MSM to be on PrEP by 2030.
3. Scenario 3 (S3): Launching an intervention in 2023 so as both high risk and medium risk MSM to be on PrEP by 2030.
4. Scenario 4 (S4): Identify the combination of interventions that could meet the HIV incidence reduction target.

The scenarios were based on three rules: a) eligible MSM did not discontinue PrEP, b) immediate discontinuation of PrEP if individuals were newly diagnosed with HIV, and c) immediate discontinuation of PrEP if MSM were no longer eligible (for example, when PrEP is only delivered to high risk MSM and MSM moved from the high risk to the medium/low risk category).

### Behavior data

Behavior data was retrieved from the EMIS-2017 national report for Cyprus (Table S2 in the supplementary material) ^19^. The risk categorization was based on a question regarding condom use with non-steady partners in the last 12 months (Table S2A). The behavior was classified in three categories of risk, i.e., high, medium, and low, and was used as input in the HIV transmission model. According to the EMIS-2017 data, 17% of the responses related to condom use during intercourse with non-steady partners could be categorized as high risk and 33.5% as medium risk.

### Sensitivity analysis

A series of univariate sensitivity analyses was undertaken to examine the impact of different model assumptions on cumulative infections under S2. These included the impact of higher/lower transmissibility while on ART with undetected viral load compared to the latent phase (85%, 99% vs. 93%), higher/lower HIV transmissibility when on PrEP (80%, 90% vs. 86%), larger/smaller MSM population with same risk structure (6000, 8000 vs. 6954), and greater/smaller proportion of MSM who are classified as high risk (10%, 20% vs. 17%).

## Results

### Model fit assessment & Status quo scenario

The status quo scenario accurately captured the overall trajectory of both HIV prevalence and HIV CoC between 2014 and 2020 (Figure 1 and Figure S2 in the supplementary material).

**Figure 1.**
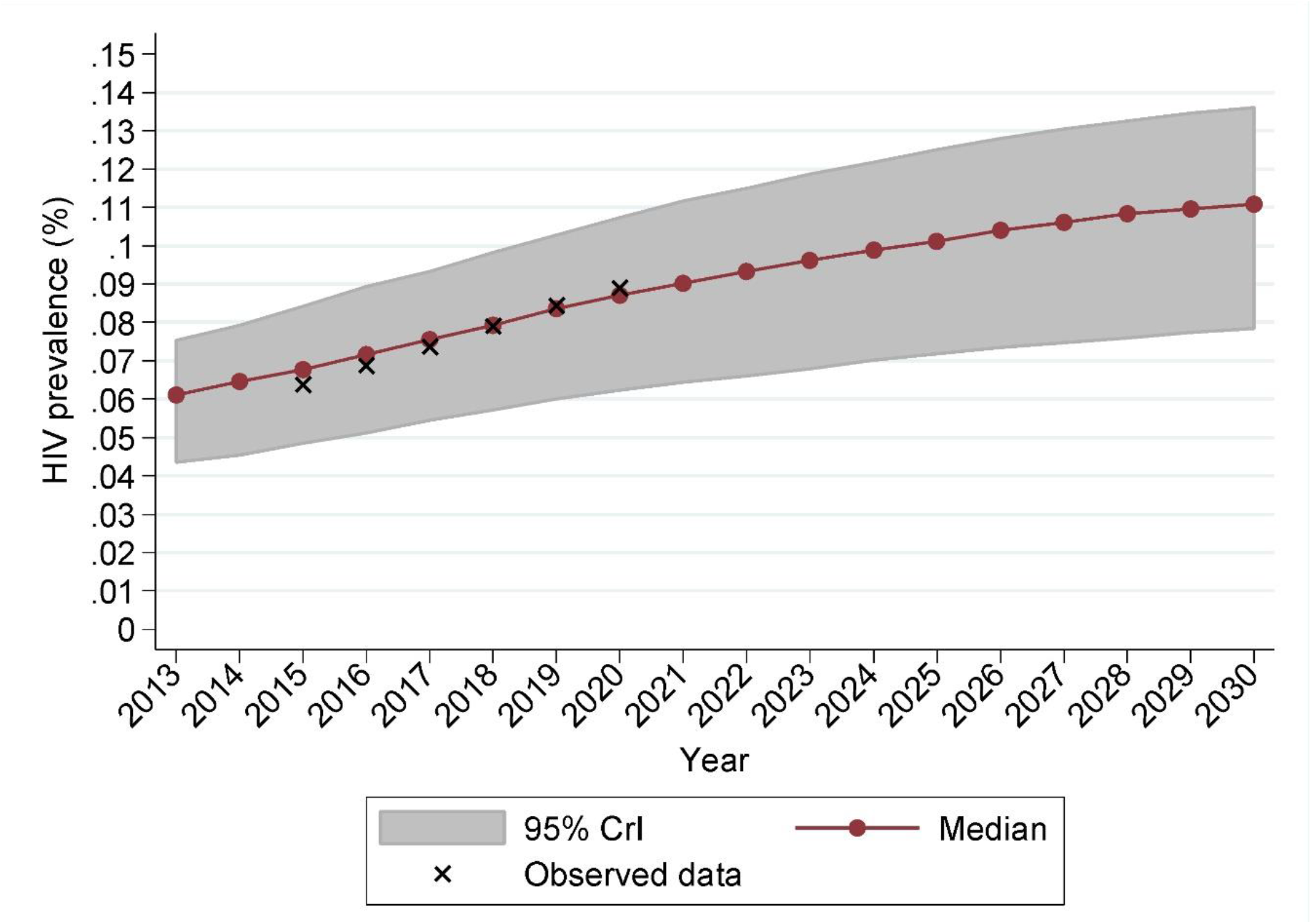
Model predictions for Human Immunodeficiency Virus (HIV) prevalence under the status quo scenario (without introduction of Pre-Exposure Prophylaxis - PrEP). The solid black line and the shaded gray error bars show the median and 95% credible intervals (95% CrI) for the model projections. For comparison, x indicate the observed HIV prevalence data.

The model estimated that the annual HIV incidence among MSM in 2020 was 3.78 (95% Credible Interval -CrI: 2.00, 6.23) per 1000 person-years. If the HIV response remains the same, the prevalence of HIV among MSM in Cyprus in 2030 is estimated at 11.1% (95% CrI: 8.0%, 14.2%) (Figure 1). This corresponds to 780 (95% CrI: 560, 995) HIV positive MSM in 2030. The relative increase in the HIV positive MSM population between 2023 and 2030 is 17.8% (95% CrI: 12.5%, 25.4%).

Under the status quo scenario, Cyprus will be very close to meeting the 95-95-95 CoC targets for MSM in 2030 (Figure S3 in the supplementary material). Specifically, at the end of 2030, 94.3% of MSM would know their status, 93.2% of them would be on ART, and 92.5% of people on treatment would be virally suppressed. However, despite the high CoC, the model suggests that the target of 90% HIV incidence reduction would not be reached: HIV incidence would decrease by 48.6% (95% CrI: 25.3%, 69.6%) in 2030 compared to 2015 (Figure 2). The cumulative HIV incidence under the status quo scenario in 2023-2030 would be 140 new infections (95% CrI: 100, 180).

**Figure 2.**
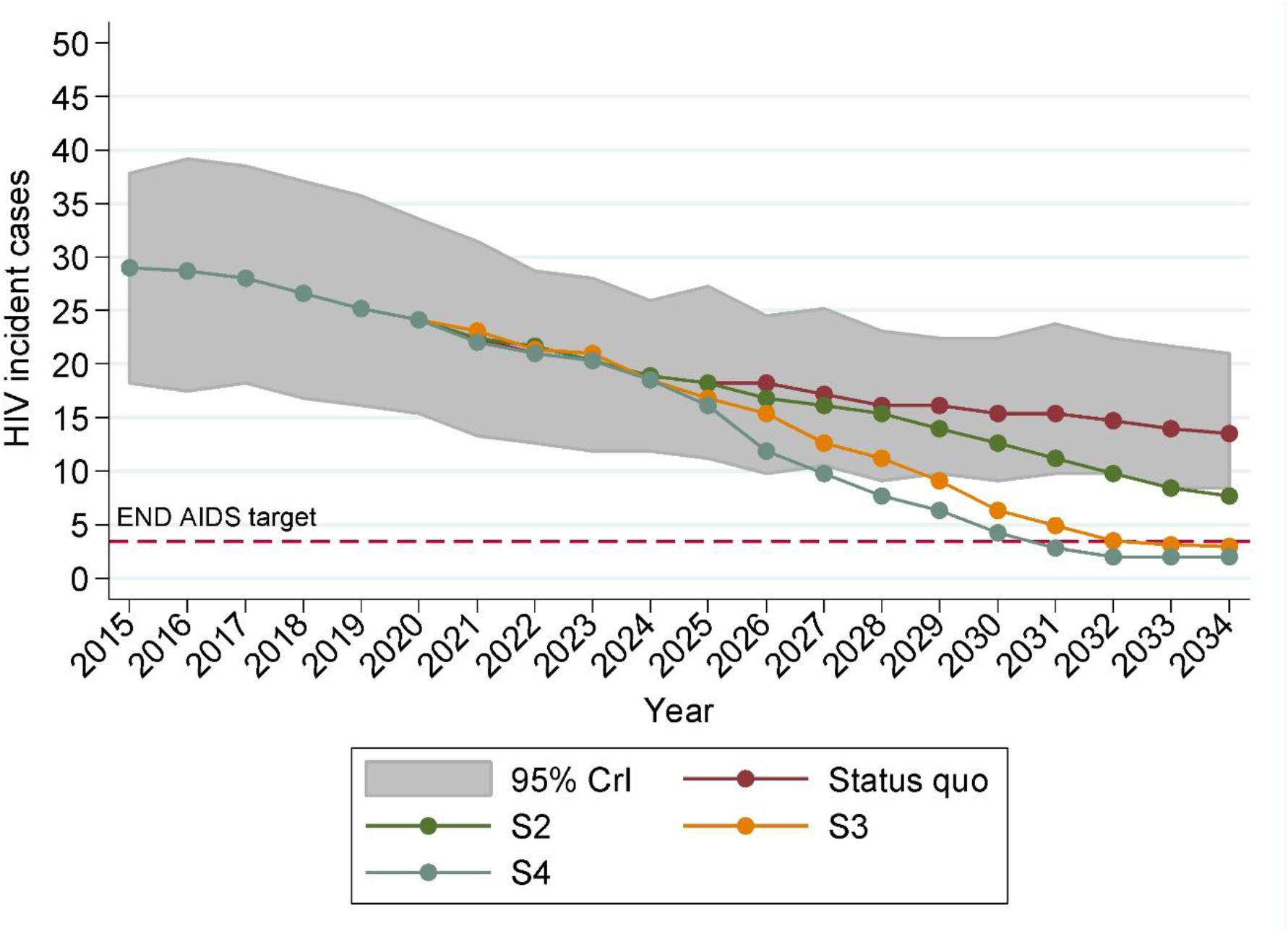
Model predictions for the Human Immunodeficiency Virus (HIV) incidence for the four different scenarios (S1: status quo scenario; S2: Launching an intervention in 2023 so as all of the high risk MSM to be on PrEP by 2030; S3: Launching an intervention in 2023 so as both high risk and medium MSM to be on PrEP by 2030; S4: Identify the combination of interventions that could meet the 90% HIV incidence reduction target by 2030). The shaded gray area bars show the 95% credible intervals (95% CrI) of the status quo scenario.

**Figure 3.**
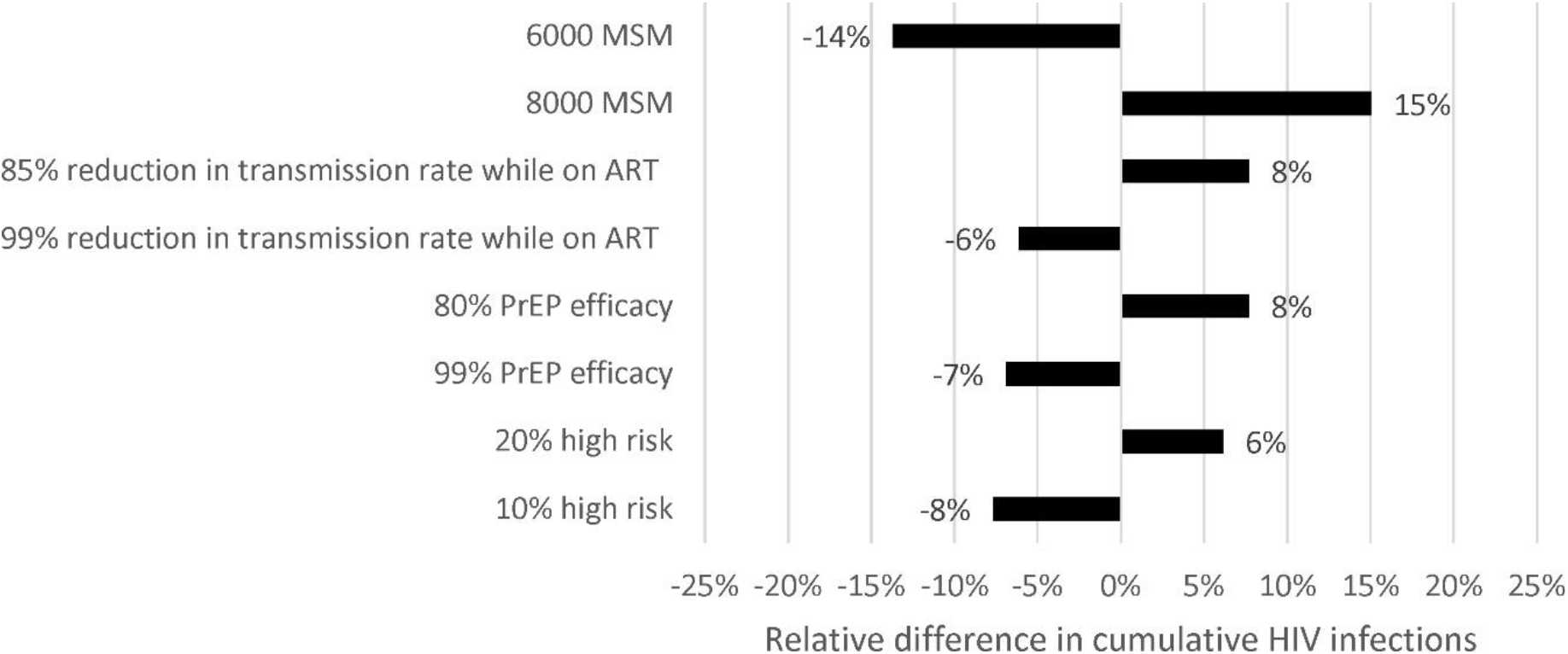
Results of one-way sensitivity analysis showing the relative difference in the cumulative number of new infections compared to the base parameter values in Table 1 under Scenario 2 (e.g., Launching an intervention in 2023 so as all of the high risk MSM to be on PrEP by 2030). A value of zero describes no change from the projected cumulative incidence. A positive or a negative value means that the projected incidence is higher or lower, respectively, compared to that projected under the baseline scenario. Abbreviations: ART, antiretroviral treatment; HIV, Human Immunodeficiency Virus; PrEP, Pre-Exposure Prophylaxis; MSM, Men who have Sex with Men.

### PrEP intervention

#### Only to high risk MSM

Starting a PrEP intervention only for high risk MSM, without other measures, would have marginal effect, since it would reduce HIV incidence by 58% (95% CrI: 39.3%, 79%) in 2030 compared to 2015 (Figure 2). Delivering PrEP only to high risk MSM would avert only 11 more HIV cases by 2030 compared to the status quo scenario, due to the small size of the high risk population in Cyprus (Figure 2).

#### High and medium risk MSM

Delivering PrEP both to high and medium risk MSM would reduce HIV incidence by 79% in 2030 compared to 2015 (95% CrI: 62.6, 90.6%) (Figure 2). The above-mentioned intervention would avert 29 more HIV cases by 2030 compared to the status quo. Under this scenario, the 90% HIV incidence reduction target would be achieved by 2032 (Figure 2). The proportion of simulations that reached the HIV incidence reduction target in 2030 was 11%.

#### HIV incidence reduction of 90%

To meet the 90% HIV reduction target in MSM by 2030, except for PrEP, interventions to reduce risky behaviors are also required. More specifically, PrEP should be expanded to both high and medium risk MSM and, after 2025, behavioral interventions should be developed and implemented so as high risk MSM gradually move to the medium risk category (Figure 2, and Figures S4 and S5 in the supplementary material).

### Sensitivity analysis

The sensitivity analysis showed that the uncertainty in the number of MSM and in the efficacy of ART and PrEP are the pivotal factors that determine the estimated number of cumulative HIV infections. Specifically, if the MSM population was larger (8000 instead of 6954), the cumulative HIV incidence would increase by 15%. On the contrary, if the size of the MSM population was smaller (6000 instead of 6954), the cumulative number of new HIV infections would decrease by 14%. Lower efficacy of ART (85% reduction in HIV transmission instead of 93%) or of PrEP (85% reduction in HIV transmission instead of 86%) would increase the expected number of new HIV infections between 2023-2030 by 8%. Similarly, a larger high risk MSM population would result in a 6% higher cumulative HIV incidence between 2023-2030.

## Discussion

Our results show that the immediate introduction of PrEP to both high and medium risk MSM in Cyprus in 2023 is a prerequisite for reaching the goal of reducing HIV incidence by 90% in 2030. Moreover, after 2025, behavioral interventions are also needed to help high risk MSM move gradually to the medium risk group.

PrEP is highly effective on preventing HIV infection and crucial for reaching the target of 90% reduction in HIV infections by 2030. However, the EMIS-2017 reported that only 44.8% of the responders in Cyprus were in favor of using PrEP if it becomes available (29% and 15.8% answered very likely and quite likely, respectively) ^17,19^. Οur model underlined the importance of running also awareness campaigns in order to promote the use of PrEP by all high and medium risk MSM. These interventions would also help overcome barriers for PrEP uptake such as fear of side-effects, inaccurate risk perception, and stigma ^12,20^. It should be also noted that the efficiency of a PrEP intervention is highly sensitive to the target population. If PrEP intervention was limited only to high risk MSM, its efficiency would be optimized but, at the same time, its public health impact would be marginal due to the small size of the high risk MSM group. On the contrary, if the eligibility criteria of PrEP were expanded to include medium risk MSM as well, the public health impact would be greater, as our model showed, but at a significantly higher cost. Thus, before defining the eligibility criteria for PrEP use, decision makers should consider the trade-off between the public health impact of PrEP and its cost.

Except from PrEP delivery, our analysis calls for behavioral interventions as well. The value of behavioral interventions to achieving the HIV incidence reduction target has been shown in other studies as well ^8,9^. Our sensitivity analysis also showed that lower effectiveness of PrEP, as a consequence of lower adherence or of an increase in risky behaviors, would make HIV incidence reduction an even more difficult target to reach. Therefore, in Cyprus, by only delivering PrEP without accompanying it with interventions to gradually reduce the high-risk population, the chances to meet the HIV incidence reduction target by 2030 are very low (11% chance to reach the target). Repeated studies should also be implemented to accurately monitor risky behavioral patterns in the MSM population.

This work has strengths and weaknesses. The strength of our model is that is based on reliably epidemiological parameters since it used the highest quality data that were available for the country. As with any modelling study, there are also limitations. First, the model ignores the impact of social networks on HIV transmission and assumes that the population is totally mixed. Second, our model was calibrated based on prevalence estimations from the ECDC tool. However, the ECDC tool is a well-known instrument that is used widely in Europe. Third, the model does not distinguish infections due to injecting drugs or sexual transmission. PrEP may be not effective for transmissions through drug injection. Fourth, our results are based on the fact that the population eligible for PrEP is correctly identified; identifying the high risk group is vital for the success of a PrEP delivery program. Additionally, PrEP users whose risk status changes (from high to low risk) may be unwilling to discontinue PrEP. Inappropriate PrEP use would increase the cost of the intervention.

## Conclusions

In conclusion, reaching the 95-95-95 CoC targets is not enough to meet the reduction of HIV incidence by 90% by 2030 in Cyprus. To end the AIDS epidemic among MSM, PrEP should be introduced in Cyprus in 2023 and distributed to both the high and medium risk MSM population. Behavioral interventions are also needed to reduce the proportion of high risk MSM. Our results, which are the work of a close collaboration between modelers, epidemiologists, and clinicians, could guide the Cypriot policymakers to make evidence-based decisions about PrEP roll-out to MSM. It should be noted, however, that the implementation of a national HIV strategy is a dynamic procedure. In order to be continuously on track to reach the HIV incidence reduction target, our mathematical model should be fed with the most up-to-date data and continually rerun.

## Data Availability

All data produced in the present study are available upon reasonable request to the authors

