## Supplement for "Modeling the HIV epidemic in MSM in Cyprus: Reaching only the 95-95-95 cascade of care targets fails to reduce HIV incidence by 90% in 2030"

### 1. Schematic outline of the mathematical model for HIV transmission

**Figure S1.** Schematic outline of the mathematical model for HIV transmission (A) and behavioral states (B) among Men who have Sex with Men (MSM). MSM begin as susceptible to infection. Once infected, they progress to the infected but undiagnosed compartment and then to the infected and diagnosed compartment. Diagnosed cases may enter the healthcare system and start antiretroviral treatment (ART). Individuals who achieve virological response have a lower probability of transmitting human immunodeficiency virus (HIV). Every year, some MSM could be lost to follow-up and return to the diagnosed but non-linked to care status. Finally, individuals can cycle from low to high risk. PrEP stands for Pre-Exposure Prophylaxis.

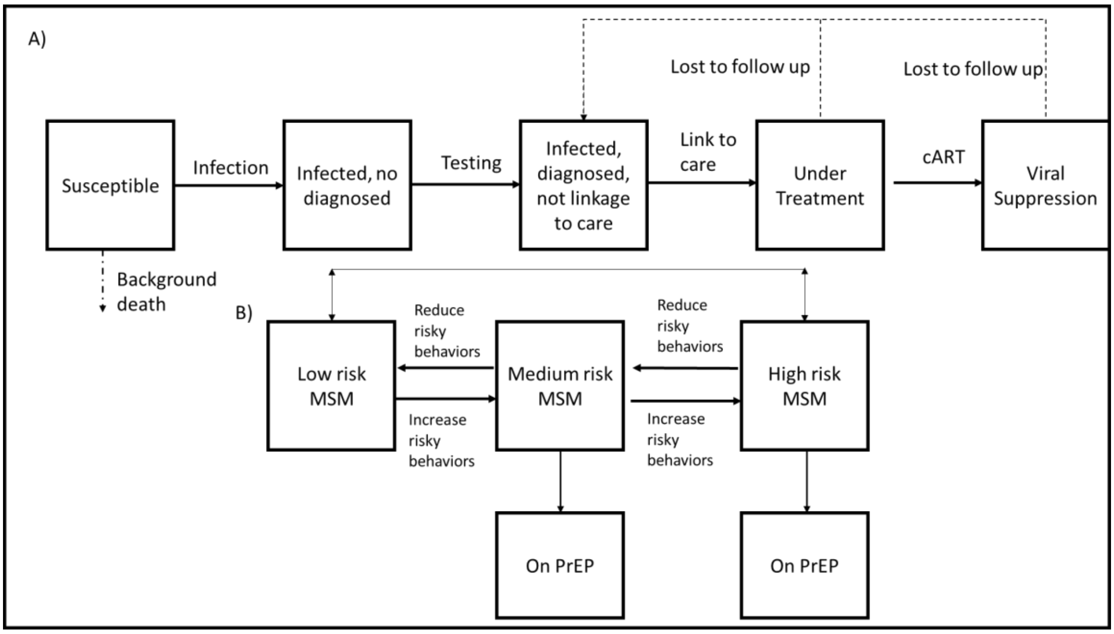

#### 2. Model type

Our analysis employed a discrete-time, stochastic, individual-based model (IBM). IBM simulates the trajectories of infected people at an individual level. These models possess some inherent randomness due to their methodology. The model examines if a pseudo-individual changes state (e.g., from susceptible to infected) using random numbers. First, the model estimates the probability of moving from one stage to the next (e.g., from susceptible to infected). Then, for each pseudo-individual, a random number from a uniform distribution (0,1) is drawn. If that random number (e.g., 0.3) is smaller than the estimated probability of changing stage (e.g., 0.4), the pseudo-individual changes stage and vice versa. For example, regarding the transmission from susceptible to infected, if the risk of infection is estimated at 20%, all individuals with corresponding random numbers in the range of 0-0.2 are assumed to get infected. As the outcome of each run depends on chance, every simulation leads to slightly different results. Uncertainty comes from a single set of parameters but across multiple simulations with randomness included. For that reason, results over all simulations are pooled and the median along a range is normally presented (stochastic variability). To assure reliability of the results, several runs should be conducted; if the number of runs is small, extreme results from simulations could significantly affect the pooled estimates. For more details regarding the IBM models, please look at [1].

#### 3. Model calibration to epidemiological and clinical data

Fitting a model to empirical data (calibration procedure) improves the confidence that the simulation model provides a realistic and accurate estimate of the outcome for health policy decisions. Thus, we used a detailed calibration procedure for our model. More specifically, we calibrated the model against reported HIV prevalence, annual HIV diagnoses, and ART initiations among MSM over time. The following sections show the methodology and the comparisons for each calibration point.

The simulations were performed in the low-level programming language C++ (Dev-C++ v.5.11) and the graphs were produced in Stata 16.1.

#### 4. Calibration estimates

Epidemiological estimates for the calibration (Table S1) were retrieved from the ECDC (European Centre for Disease Prevention and Control) tool [2] following its application to data from the HIV reporting system in the Republic of Cyprus until 2020 (last estimates from the tool were obtained in January 2022).

**Table S1.** Calibration parameters.

| Year | HIV prevalence in MSM (% of MSM who are infected with HIV) | % of MSM with HIV infection who are diagnosed | % of the diagnosed MSM with HIV who are on ART | % of MSM with HIV who are on ART with undetectable VL (<1000 copies/ml) |
|---|---|---|---|---|
| 2015 | 6.37% | 76.9% | 80% | 89.8% |
| 2016 | 6.88% | 81.3% | 88% | 95.6% |
| 2017 | 7.37% | 84.5% | 79% | 97.5% |
| 2018 | 7.91% | 86,7% | 90% | 95.4% |
| 2019 | 8.44% | 89.5% | 86% | 92.1% |
| 2020 | 9.01% | 92.2% | 88% | 92.9% |

Note: MSM: Men who have Sex with Men; ART: antiretroviral treatment; VL: viral load

#### 5. Goodness-of-fit metric (GoF)

The GoF metric measures the accuracy of the ability of the model to predict the calibration points. Our model used the weighted least square method for that purpose. The GoF is estimated as follows:

$$GoF_{overall}=\sum (w_{prevalence*} (Modelled\ prevalence\text{-}observed\ prevalence)^2 + w_{diagnosis*} (Modelled\ diagnoses\text{-}observed\ diagnoses)^2 + w_{treatment*} (Modelled\ treatments\text{-}observed\ treatments)^2)$$

where w denotes weights.

The weighting factors leave preferences on the set of the evaluated targets and are judged according to the relative importance of those targets. In this modelling work, we ranked each target equally important with respect to the fit of the model. Thus, we assigned a weight of 0.33 to all targets. Smaller values of the GoF metric indicate a better fit to the observed data.

#### 6. Modelling the increased infectivity during the acute phase

The model has been developed using one-year time steps. We assumed that the probability of infection was homogeneously distributed across the months of a year. Due to higher viremia during the early stages of HIV infection, recently HIV-infected MSM are more likely to transmit the virus than those who are longer-term infected [3–6]. To account for the increased infectivity of recently HIV-infected MSM, the model assumes that a) MSM are 14.5 times more infectious during the period of recent HIV infection than MSM in later phases of HIV infection, and b) this high viremia period lasts for the 3 first months after HIV acquisition. Based on these assumptions, we implemented the following calculation:

*3/12 (3 out of the 12 months) *14.5 (Cofactor increase in HIV transmission during the period of recent infection) + 9/12 (9 out of 12 months) *1 (Cofactor increase in HIV transmission after the period of recent infection) = 4.375*

Based on the above calculation, an HIV-infected MSM in the 1st year of his infection had a 4.375 times higher probability of transmitting HIV than an HIV-infected MSM who had acquired the virus more than a year ago.

#### 7. European MSM Internet Survey - EMIS-2017 data

European Men-Who-Have-Sex-With-Men Internet Survey (EMIS-2017) was an international internet-based survey for MSM. It was conducted simultaneously in 50 countries including Cyprus from 18 October 2017 to 31 January 2018. The participants were men living in Europe or in a limited number of countries outside Europe, at or over the age of homosexual consent, and who were sexually attracted to men and/or had sex with men [7].

The aim of EMIS-2017 was to measure: (a) sexual health outcomes, (b) risk and precaution behaviors, (c) health promotion needs, and (d) coverage/uptake of interventions. National reports are available at the EMIS-2017 website (www.emis2017.eu) and relevant studies were published providing an overview of the characteristics of MSM while identifying needs for policy planning. The findings of EMIS-2017 in Cyprus were based on 307 eligible questionnaires of participants with a median age of 35 years (interquartile range 28-44 years) [8].

**Table S2.** Risk groups of Men who have Sex with Men (MSM) in Cyprus according to their behavior and willingness to receive Pre-Exposure Prophylaxis (PrEP) – data from the European Men-Who-Have-Sex-With-Men Internet Survey (EMIS-2017).

| | | **n** | **Percentage (%)** | **Risk group** |
|---|---|---|---|---|
| **A** | How often were condoms used when you had intercourse with non-steady male partners? | | | |
| | Never | 14 | 7.7 | High |
| | Seldom | 6 | 3.3 | High |
| | Sometimes | 11 | 6.0 | High |
| | Mostly | 61 | 33.5 | Medium |
| | Always | 90 | 49.4 | Low |
| | Total | 182 | 100 | |
| **B** | If PrEP was available and affordable to you, how likely would you be to use it? | | | |
| | Very unlikely | 61 | 22.9 | |
| | Quite unlikely | 14 | 5.3 | |
| | Not sure | 69 | 25.9 | |
| | Quite likely | 42 | 15.8 | |
| | Very likely | 77 | 29.0 | |
| | Not answered | 3 | 1.1 | |
| | Total | 266 | 100 | |

#### 8. Model fit assessment

**Figure S2.** Model predictions for the cascade of care (CoC) among men who have sex with men in Cyprus in (A) 2019 and (B) 2020 under the status quo scenario (without Pre-Exposure Prophylaxis - PrEP). ART: antiretroviral treatment.

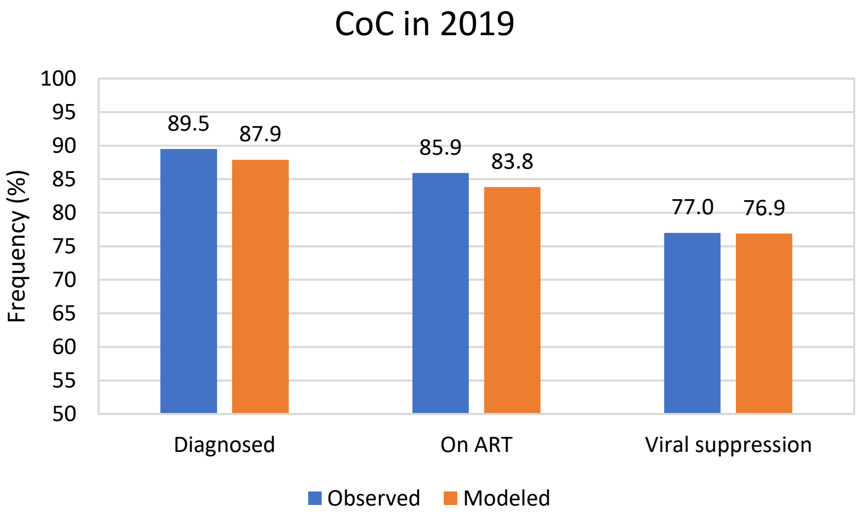

(A)

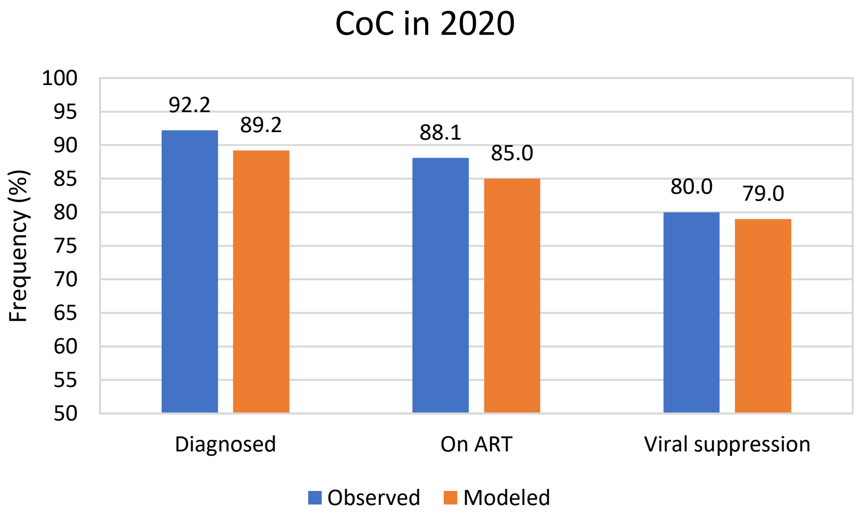

(B)

**Figure S3:** Model predictions for the cascade of care (CoC) among men who have sex with men in Cyprus in 2030 under the status quo scenario (without Pre-Exposure Prophylaxis - PrEP). ART: antiretroviral treatment.

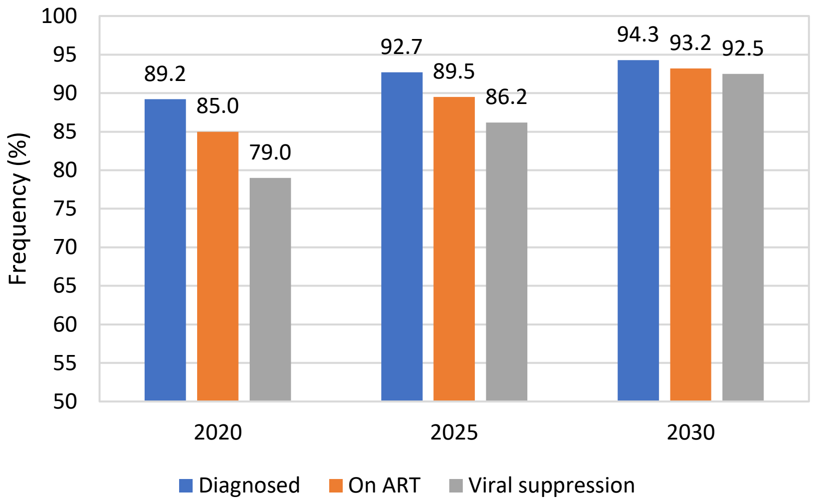

#### 9. Detailed projections with credible intervals

In the main text, we included credible intervals (CrIs) only for the status quo scenario, otherwise the figures would be hard to read. Thus, we present below the CrIs of all scenarios compared to the status quo (without Pre-Exposure Prophylaxis – PrEP).

**Figure S4.** Model predictions including credible intervals (CrIs) for HIV prevalence - status quo scenario (without Pre-Exposure Prophylaxis - PrEP) and the scenario where only high risk Men who have Sex with Men (MSM) are eligible for PrEP.

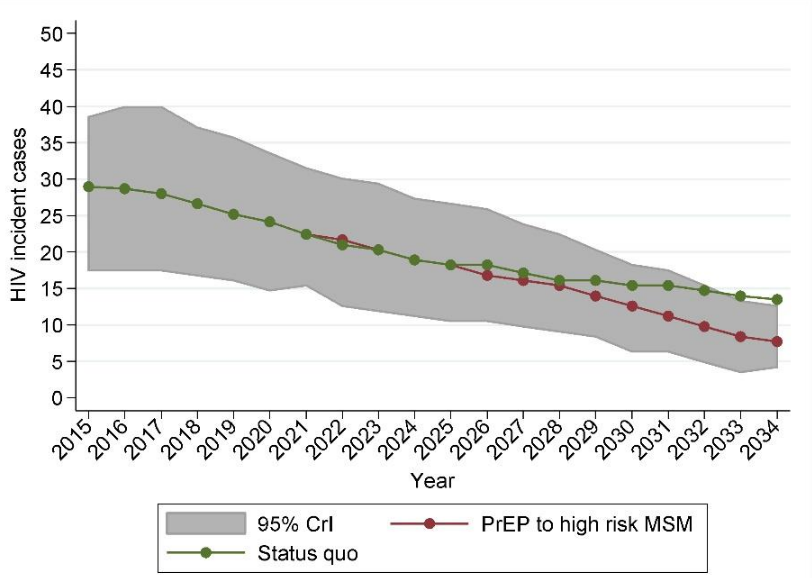

**Figure S5.** Model predictions including credible intervals (CrIs) for HIV prevalence - status quo scenario (without Pre-Exposure Prophylaxis - PrEP) and the scenario where high and medium risk Men who have Sex with Men (MSM) are eligible for PrEP.

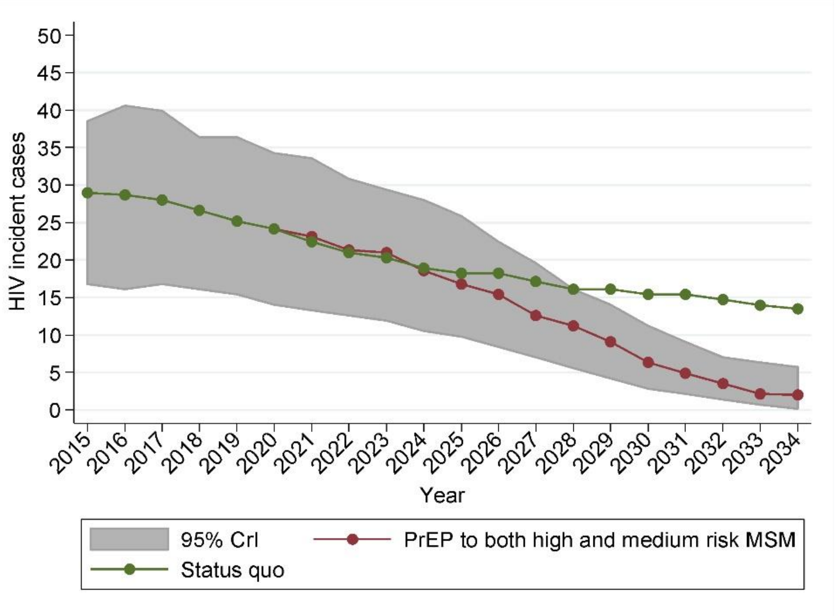
